# 2D versus 3D artificial intelligence-driven segmentations of airway alterations in cystic fibrosis: which one is better?

**DOI:** 10.1101/2023.11.18.23298712

**Authors:** Imene Hadj Bouzid, Ilyes Benlala, Gael Dournes

## Abstract

**Purpose or Learning Objective:** Artificial intelligence (AI) with convolutional neural network allows fully automated detection and segmentation of bronchial changes on CT-scans of cystic fibrosis (CF). However, the superiority of two-dimensional (2D) versus three-dimensional (3D) architectures remains to be explored.

**Method or Background:** CT-scans from fifty CF patients were retrospectively included at two CF reference centers. The nnUnet model was implemented in both 2D and 3D, and trained to segment five structural alterations: bronchiectasis, wall thickening, mucus plugs, bronchiolar impactions and consolidations. A semantic validation was done by using fifty CTs with a five-fold cross validation strategy, by comparing normalized Dice-Sorensen coefficient (DSC) between 2D and 3D architectures, with manual segmentations as Gold Standard.

**Results or Findings:** The 3D nnUnet was found able to segment the five CF main hallmarks such as bronchiectasis, wall thickening, mucus plugs, bronchiolar impactions and consolidations. Metrics obtained with the 3D architecture were superior for mucus plugs, bronchiolar impactions and consolidations (p<0.001) but not significantly different for bronchiectasis and wall thickening (p>0.05).

**Conclusion:** AI with the 3D-nnUnet model can perform fully automated segmentation of CF-related structural hallmarks on CT scans, and overcome 2D implementation. Non-invasive, holistic 3D quantifications are allowed for promising next clinical applications.

## INTRODUCTION

Artificial intelligence (AI) with deep learning (DL) is a recent breakthrough in medical imaging, which is changing the landscape of diagnostic tools available [1]. Beyond human capacities [2], DL algorithms with convolutional neural networks (CNN) can perform computer vision tasks such as detection and segmentation of disease-related abnormalities on CT scans in a reproducible and rapid manner, which could prove useful for quantitative imaging purpose [3]. In the field of airway imaging using CT, there is a growing need for non-invasive characterization and quantification of the lung structural abnormalities, to assess the disease severity and the longitudinal modifications over time [4]. For instance, cystic fibrosis (CF) remains one of the most frequent genetic disorders in Caucasians, affecting up to 1 every 2500 children [5]. The lung is the most affected organ, where dysfunction of the CFTR protein within airways is responsible for an increased production of thick and sticky mucus, leading to pulmonary exacerbations and death. However, effective CFTR modulator treatments have recently emerged and can dramatically modify the disease course with an improvement of both clinical symptoms and lung structural damages. Therefore, availability of non-invasive biomarkers for allowing objective and reproducible quantitative assessments of the lung disease process may be desirable to help the clinicians for their disease management [6].

CT of the lung in CF has been demonstrated useful to complete this task [7]. Indeed, CT can demonstrate the main structural hallmarks of CF such as bronchiectasis, airway wall thickening, mucus plugins and consolidations [8], in a non-invasive manner. Several visual scoring systems have been proposed to correlate with the lung disease severity. Nevertheless, these visual methods are subjective and may lack reproducibility between different readers [9]. Also, these systems are using categories to summarize the extent of structural damages, which may lack sensitivity, especially for longitudinal application. Recently, AI has been proven to allow rapid and reliable depiction of CF hallmarks in a fully automated manner [10]. This approach demonstrated the possibility to reach a holistic quantification of the 3D volumetric extent of lung lesions, over a full set of CT slices for a given patient. However, previous study was done by using 2D CNNs only, while the volume of lesions was obtained as the sum of individual 2D results. Conversely, the feasibility of a genuine 3D approach was not assessed. On the one hand, 3D segmentation methodologies capitalize on the comprehensive spatial context of volumetric data, ensuring consistent segmentations that accurately capture intricate three-dimensional structures. On the other hand, 2D techniques, guarantee rapid processing times and a direct application especially when considering the ubiquity of 2D medical imaging modalities.

The aim of the study was to assess the performance of a 3D-CNN model to detect and quantify CF structural abnormalities on CT scans of CF patients, and to compare the evaluation to that of its 2D-CNN counterpart. Secondary objective was also to correlate the volumetric quantifications with pulmonary function tests (PFT).

## MATERIAL AND METHODS

### Study design

The retrospective study was held between January 2020 and December 2022 at a single Institution, involving two CF reference centers dedicated to children and adults. All patients were provided written informed consent for reusing data from their medical records, after approval by the Ethic Committee of the University Hospital of Bordeaux, (Full name: “**Direction de la qualité et de la gestion des risques”; Affiliation:** University Hospital of Bordeaux; study registration number: CHUBX2020RE0267).) in compliance with French laws (https://www.legifrance.gouv.fr/jorf/id/JORFTEXT000037187498). Inclusion criteria were: age greater than 8-year-old, CF diagnosis made on sweat chloride and/or genetic test, non-contrast-enhanced CT alongside PFT performed the same day. Disease management was performed according to a standard of care. There were no exclusion criteria.

### CT of the lung

Supplemental Table E describes the CT characteristics. There were two different machine models from 2 major manufacturers, namely GE Revolution®, and Siemens Somatom Force®. The matrix was 512*512, the dose-length product ranged from 8 to 260 mGy.cm and the slice thickness from 1 to 1.25 mm. All patients were thoroughly coached in breathing techniques before each CT scan and CT at full inspiration and reconstructed with standard kernels.

#### Methodology used for labeling of CT slices

The annotation of CT slices was done in consensus between three observers of 6, 12, and 25 years of experience in thoracic imaging, who are part of a CF reference center which belongs to the European Cystic Fibrosis Society Clinical Trial Network, and with published expertise in CF scoring of lung CT and MRI[12–16].

Manual segmentation of labels was performed by using the 3D Slicer software 4.11, an open-source software. CT images were displayed with parenchymal window width and level (width, 1500 Hounsfield Unit; level -450 Hounsfield Unit)[17]. Five labels were created to represent five main hallmarks of structural alterations of CF: bronchiectasis, peribronchial thickening, bronchial mucus plugs, bronchiolar mucus plugs with the “tree-in-bud” pattern, and collapse/consolidation[18]. In this study, bronchiectasis refers to the mucus-free airway lumen dilatation, and the bronchial mucus plug was scored when a secretion filled the bronchial lumen entirely. A sixth label was also created, which corresponds to the lung parenchyma, as the total lung minus the sum of other abnormal labels. A visual agreement between the three observers of more than 80% in the visible spatial extent of true-positive findings was necessary.

### AI framework

For the segmentation of CF lesions, a detailed semantic segmentation process was initiated. CT volumes from 50 patients were incorporated to structure the training and validation datasets. Slice-by-slice manual segmentations were carried out to delineate five labels: bronchiectasis, bronchial wall thickening, bronchial mucus, bronchiolitar impaction with the “tree-in-bud” pattern, and consolidation. For cross-validation purposes, the CTs were partitioned into five groups, with each group consisting of 10 randomly assigned patients.

The nnU-Net architecture, in both 2D and 3D implementations, was utilized for training, with more details provided in the supplemental materials (Supplemental Method). A dynamic loss function was implemented, initially emphasizing the Dice Coefficient. Over epochs, the focus shifted to the bottom 50% of predictions (TopK). This approach guaranteed foundational segmentation accuracy in the initial stages and refined precision for challenging areas in subsequent training phases [11].

A dual-modality input was employed for nnU-Net. The primary modality showcased the hole CT scan, while the secondary displayed only the inner region of the lung parenchyma. This dual-input strategy aimed to direct the network’s attention to essential intra-lung regions, avoiding extraneous external components. The methodology is depicted in Figure 2.

**Figure 1.**
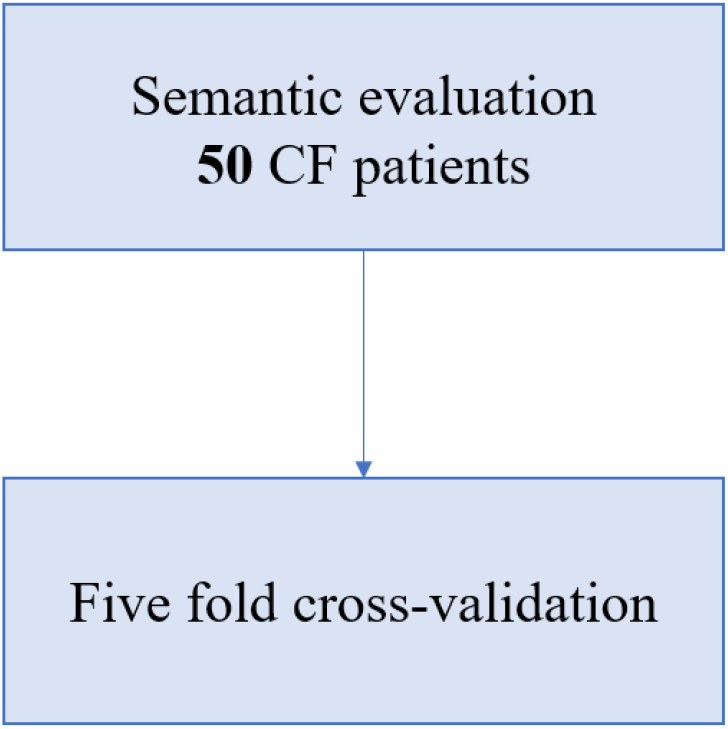
Study flow chart. CF=cystic fibrosis; CT=computed tomography; PFT = pulmonary function test.

**Figure 2.**
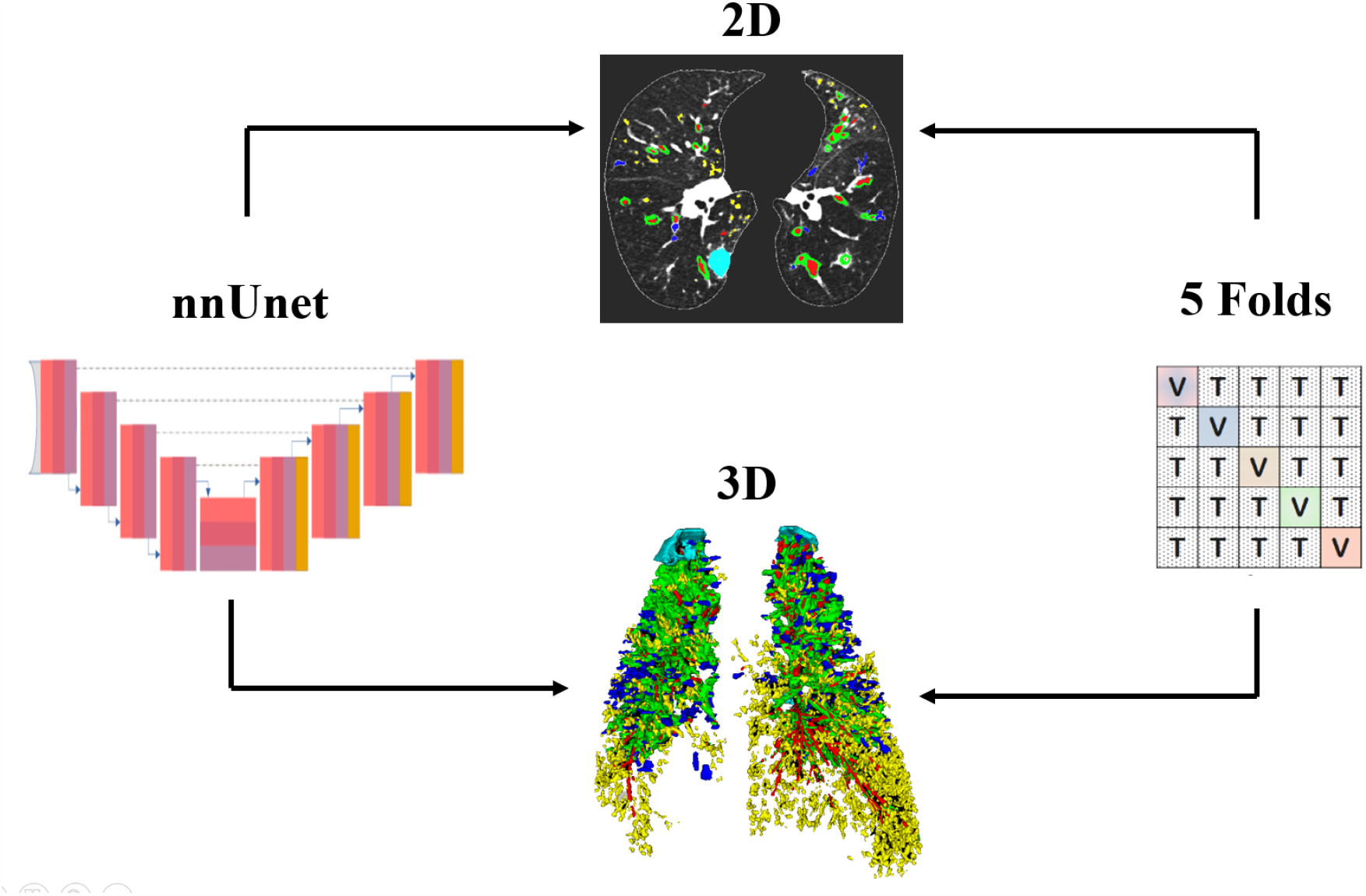
Study methodology. The convolution neural network is nnUnet ; two dimensional (2D) and three dimensional (3D) architectures were trained with databases consisting of CT slices and CT volume. The validation was conducted using a 5-fold cross-validation : The data set is divided into 5 equally folds, the model is trained on 4 of these folds (T) and tested on the remaining one (V).

### Software

The 2D and 3D nnUnet were trained on a system operating on Ubuntu 18.04. The environment was set up with Python 3.9, using PyTorch 2.0 and CUDA 11. The system’s hardware was anchored by an Intel Xeon Gold 5217 CPU, featuring 2 physical processors and a total of 32 threads, 30 of which were allocated exclusively to nnU-Net during training. The system was bolstered by 200GB of RAM and employed a Quadro RTX 8000 GPU with 48GB of VRAM.

### Performance metrics

To evaluate the precision of the segmentation models related to CF lesions, reliance was primarily placed on the Dice Coefficient (DSC). This metric is effective in gauging the overlap between the predicted and actual regions. However, its efficacy diminishes when addressing small structures [12]. This limitation led to the consideration of the complementary attributes of the Normalized Surface Distance (NSD). NSD excels at recognizing instances where predictions are in close proximity but not perfectly overlapping with the true lesions, underscoring its clinical relevance.

Precision was employed to gauge the correctness of positive detections, while Recall was used to ensure comprehensive identification of notable structures. Additionally, the Area Under the Curve (AUC) was used as a significant metric, capturing the model’s capability to distinguish between classes effectively. Confusion matrices were also incorporated into the evaluation metrics, providing a clear and concise visualization of prediction misclassifications.

### Interpretability

To further understand the inner workings of nnUnet, the visualization technique known as Gradient-weighted Class Activation Mapping (Grad-CAM [13]) was utilized. This tool produced a heatmap highlighting regions of the image considered crucial by the model. Additionally, exploration was made into a Bayesian neural network methodology [14]. The model was examined through five distinct iterations to ascertain its intrinsic consistency and gain insight into its predictive confidence.

### Statistical tests

Comparisons of continuous data were done using the Mann Whitney test, and categorical data with the Chi-square test. A p-value inferior to 0.05 was considered significant.

## RESULTS

### Performance metrics

Regarding internal validation, it was observed that both models demonstrated comparable performance for bronchiectasis, wall thickening, and consolidations, Indeed, nnUnet 2D achieved mean scores of 0.77 (±0.056), 0.64 (±0.044), and 0.61 (±0.298), respectively, while its 3D version garnered evaluations of 0.81 (±0.043), 0.65 (±0.034), and 0.60 (±0.333), respectively.

However, the 3D version excelled over its 2D counterpart in detecting bronchial and bronchioliar mucus, with an improvement of 8% for the bronchial mucus and nearly 12% for the bronchiolitis mucus (Figure 3). The superior NSD performance, in comparison to Dice, suggests that models detect at the right locations, albeit without exhaustively capturing the shapes of the structures, this was particularly noticeable for wall thickening.

**Figure 3.**
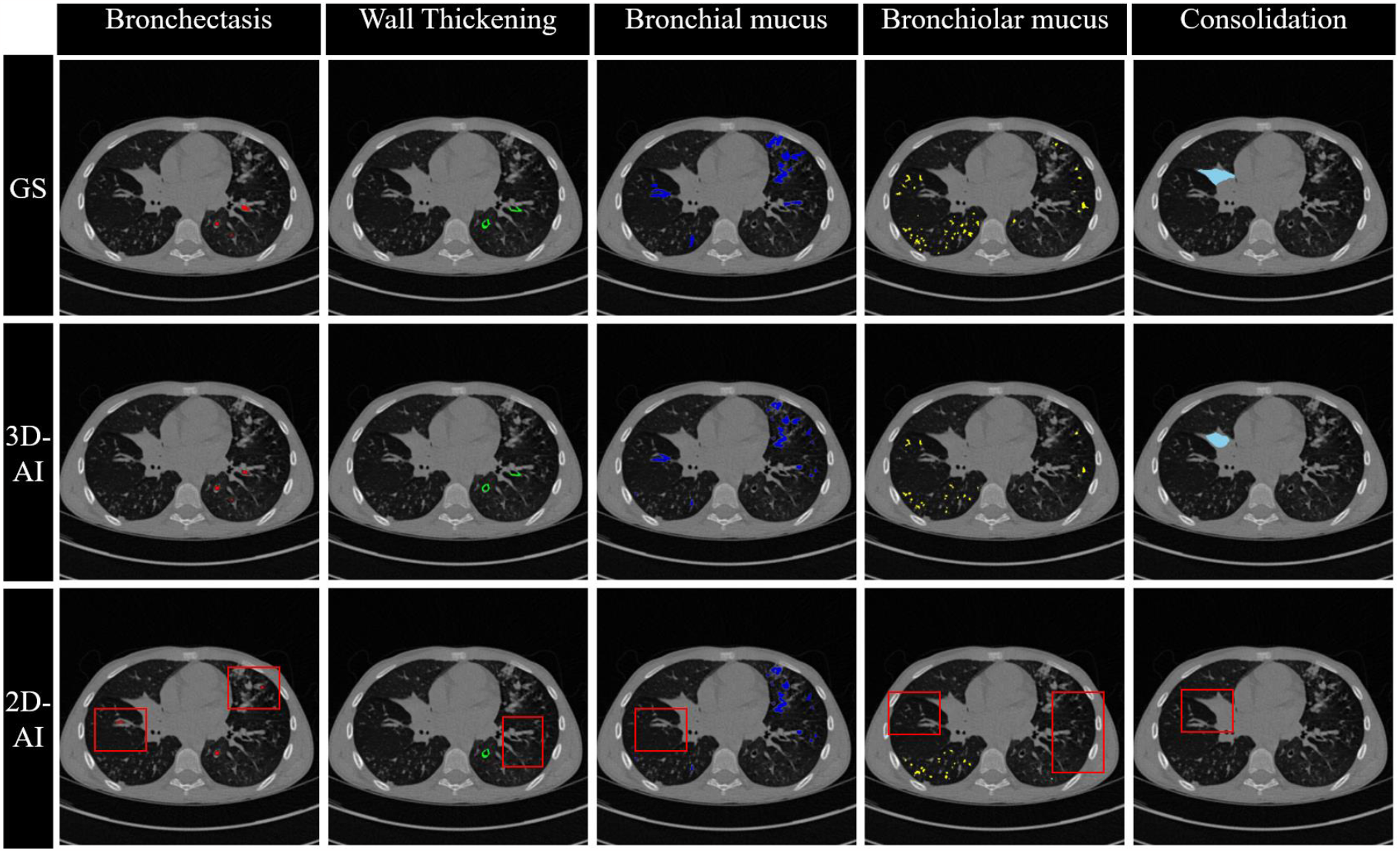
Comparison of CF lesion segmentations (Bronchiectasis, Thickening, Bronchial mucus, Bronchiolar mucus, Consolidation). The first row displays the segmentations from the Gold Standard (GS), the second row presents the segmentations predicted by nnUnet (**3D-AI**), and the third row depicts the segmentations produced by nnUnet (**2D-AI**). The red squares highlight the errors made by the 2D version across the five CF lesions. On the same slice, structures corresponding to each of the five CF labels are observed to have been omitted by the 2D nnUnet version when compared to its 3D counterpart.

Specificity and sensitivity values indicated that the models were specific, with the 3D version exhibiting enhanced sensitivity. The increase in sensitivity is evident in the detection of bronchial mucus, which improved by 10% compared to the 2D segmentation, whereas the bronchiolitis mucus saw a gain of 12% (Figure 4). Lastly, the AUC scores for the 2D segmentation ranged between 0.613 (±0.042) for bronchiolitis mucus and 0.87 (±0.034) for bronchiectasis, while the AUCs for nnUnet 3D spanned from 0.683 (±0.042) for bronchiolitis mucus to 0.886 (±0.030) for bronchiectasis. The detailed results of these evaluations are provided in the Table 1.

**Table 1.** Segmentation performances of the nnUnet 2D and 3D across the CF lesions: Bronchiectasis, Wall Thickening, Bronchial Mucus, Bronchiolitis Mucus and Consolidation.

|  | Bronchiectasis |  | Thickening |  | Bronchial Mucus |  | Bronchiolitis Mucus |  | Consolidation |  |
| --- | --- | --- | --- | --- | --- | --- | --- | --- | --- | --- |
|  | 2D | 3D | 2D | 3D | 2D | 3D | 2D | 3D | 2D | 3D |
| DICE | 0.77($\pm 0.05$) | 0.80($\pm 0.043$) | 0.63($\pm 0.04$) | 0.65($\pm 0.03$) | 0.55($\pm 0.08$) | 0.64($\pm 0.03$) | 0.28($\pm 0.05$) | 0.39($\pm 0.05$) | 0.61($\pm 0.29$) | 0.60($\pm 0.33$) |
| NSD | 0.79($\pm 0.05$) | 0.80($\pm 0.06$) | 0.81($\pm 0.05$) | 0.81($\pm 0.05$) | 0.60($\pm 0.05$) | 0.68($\pm 0.11$) | 0.44($\pm 0.05$) | 0.51($\pm 0.08$) | 0.53($\pm 0.05$) | 0.65($\pm 0.08$) |
| AUC | 0.87($\pm 0.03$) | 0.886($\pm 0.030$) | 0.81($\pm 0.04$) | 0.82($\pm 0.03$) | 0.73($\pm 0.064$) | 0.78($\pm 0.035$) | 0.61($\pm 0.03$) | 0.68($\pm 0.04$) | 0.82($\pm 0.07$) | 0.80($\pm 0.07$) |

**Figure 4.**
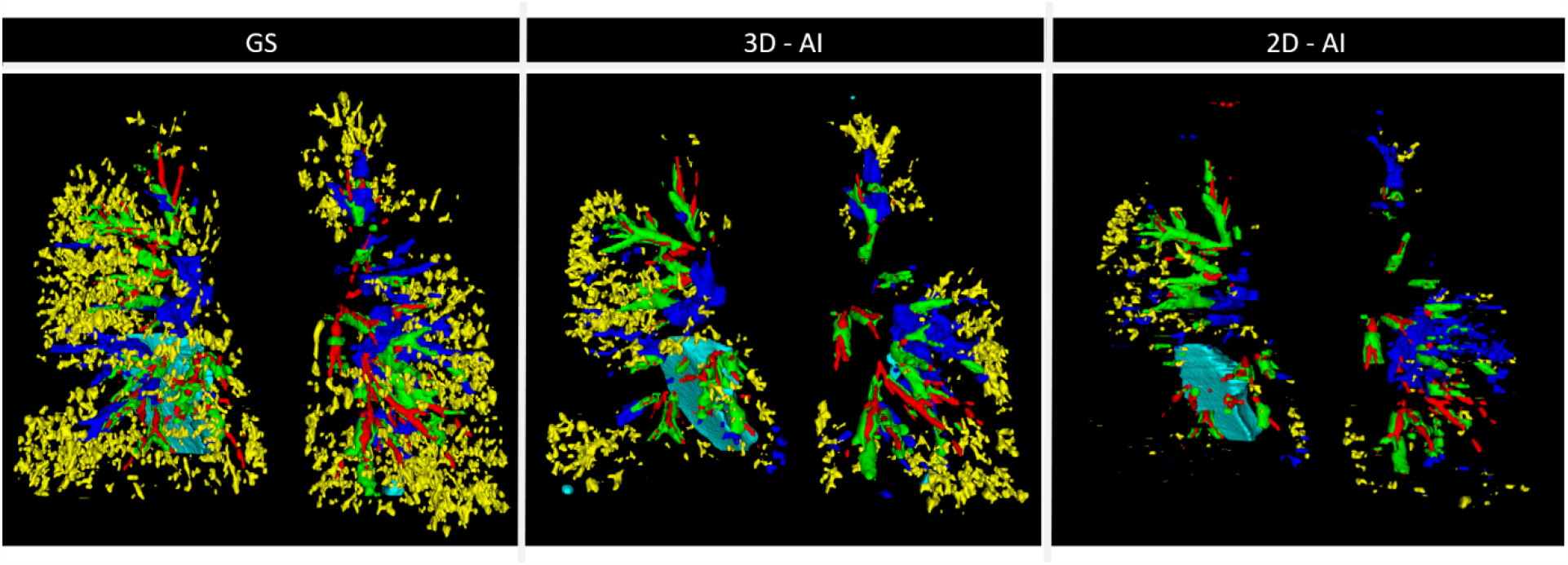
Comparison of CF lesion segmentations (Bronchiectasis, Thickening, Bronchial mucus, Bronchiolar mucus, Consolidation). The first column presents the 3D segmentations from the Gold Standard (GS), the second column represents the 3D segmentations predicted by nnUnet (**3D-AI**), and the third column displays the 3D segmentations produced by nnUnet (**2D-AI**). The 3D segmentation of CT scans from CF patient resulted in a notable increase in sensitivity, especially evident for the bronchiolitis mucus (yellow), compared to its 2D counterpart.

Confusion matrices for both 2D nnUnet (Table 2) and 3D nnUnet (Table 3) indicate that for bronchiectasis and wall thickening, there is minimal confusion with other lesions regardless of whether the segmentation is 2D or 3D. However, the two types of mucus were more often confused with each other, whereas the consolidations are mistakenly identified as wall thickening or bronchiolitis mucus. The 3D version has mitigated these confusions, enhancing scores by 3%, 6%, and 2% for bronchial mucus, bronchiolitis mucus, and consolidation respectively (p<0.01). Additionally, the standard deviations of 3D nnUnet were also reduced for all lesions collectively.

**Table 2.** Confusion matrix for 2D nnUnet. Each column represents the distribution of predictions by lesion type.

| 2D | Bronchiectasis | Thickening | Bronchial Mucus | Bronchiolar Mucus | Consolidation |
| --- | --- | --- | --- | --- | --- |
| Bronchiectasis | 0.98 (± 0.00) | 0.01(± 0.00) | 0.00<br>(± 0.00) | 0.00(± 0.00) | 0.00(± 0.00) |
| Thickening | 0.01 (± 0.00) | 0.95 (± 0.01) | 0.08 (± 0.03) | 0.01 (± 0.00) | 0.05(± 0.04) |
| Bronchial | 0.00 (± 0.00) | 0.02 (± 0.00) | 0.80 (± 0.11) | 0.15 (± 0.11) | 0.04 (± 0.03) |
| Bronchiolitis | 0.00 (± 0.00) | 0.00 (± 0.00) | 0.11 (± 0.09) | 0.83 (± 0.11) | 0.00 (± 0.00) |
| Consolidation | 0.00 (± 0.00) | 0.00 (± 0.01) | 0.00 (± 0.00) | 0.00 (± 0.00) | 0.90<br>(± 0.07) |

**Table 3.**
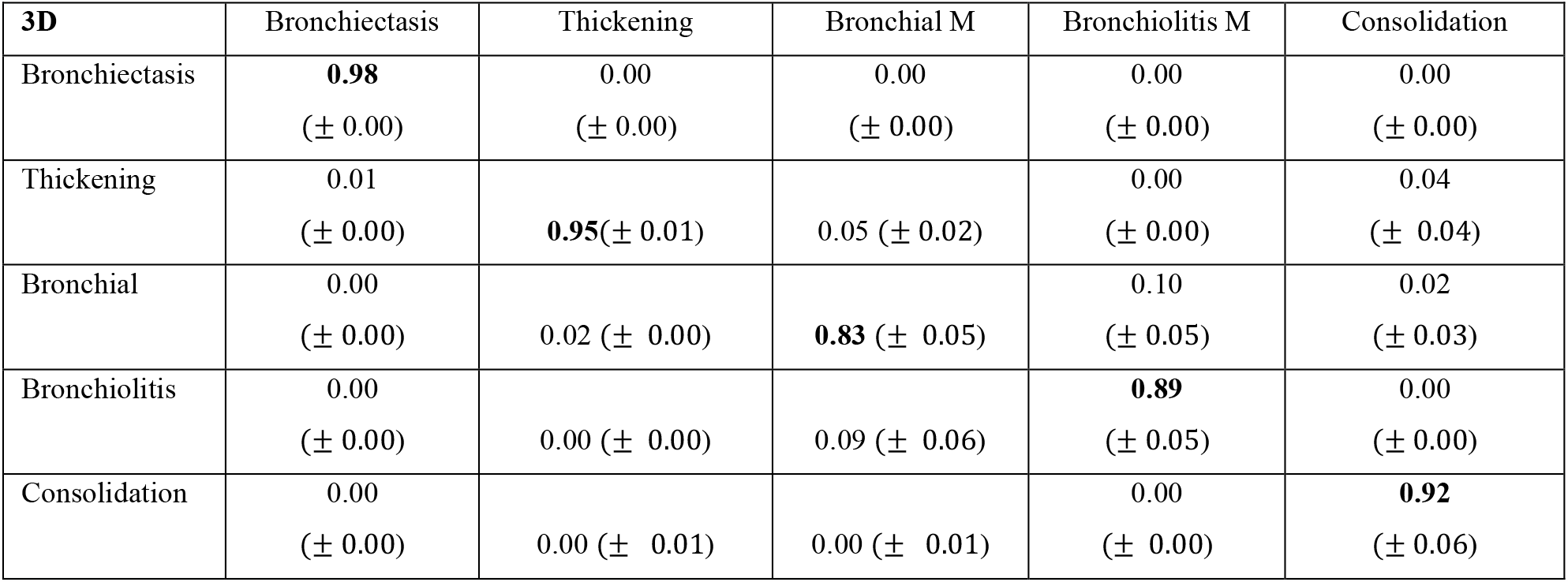
Confusion matrix for 3D nnUnet. Each column represents the distribution of predictions by lesion type.

| **3D** | Bronchiectasis | Thickening | Bronchial M | Bronchiolitis M | Consolidation |
| --- | --- | --- | --- | --- | --- |
| Bronchiectasis | **0.98**<br>(± 0.00) | 0.00<br>(± 0.00) | 0.00<br>(± 0.00) | 0.00<br>(± 0.00) | 0.00<br>(± 0.00) |
| Thickening | 0.01<br>(± 0.00) | **0.95**(± 0.01) | 0.05 (± 0.02) | 0.00<br>(± 0.00) | 0.04<br>(± 0.04) |
| Bronchial | 0.00<br>(± 0.00) | 0.02 (± 0.00) | **0.83** (± 0.05) | 0.10<br>(± 0.05) | 0.02<br>(± 0.03) |
| Bronchiolitis | 0.00<br>(± 0.00) | 0.00 (± 0.00) | 0.09 (± 0.06) | **0.89**<br>(± 0.05) | 0.00<br>(± 0.00) |
| Consolidation | 0.00<br>(± 0.00) | 0.00 (± 0.01) | 0.00 (± 0.01) | 0.00<br>(± 0.00) | **0.92**<br>(± 0.06) |

### Models’s interpretability

The feature maps obtained through the GradCAM algorithm (Figure 5) clearly demonstrate that both the 2D and 3D models focus on the regions of interest before opting for a labeling. Notably, the 2D network appears to exhibit more hesitation compared to the 3D version, especially in the case of mucus identification. Moreover, the differences estimated between the five versions of nnUnet in both 2D and 3D versions deviate by only 10^−4^ (Table 4), further corroborating the robustness of the network in its predictions.

**Table 4.** Uncertainty evaluation of the 2D and 3D nnUnet across the 5 five CF lesions : Bronchiectasis, Wall bronchiectasis, Bronchial Mucus, Bronchiolitis Mucus, Consolidation.

| Uncertainty | Bronchiectasis | Thickening | Bronchial Mucus | Bronchiolitis Mucus | Consolidation |
| --- | --- | --- | --- | --- | --- |
| **2D** | 0.00 (± 0.00) | 0.00 (± 0.00) | 0.00 (± 0.00) | 0.00 (± 0.00) | 0.00 (± 0.00) |
| **3D** | 0.00 (± 0.00) | 0.00 (± 0.00) | 0.00 (± 0.00) | 0.00 (± 0.00) | 0.00 (± 0.00) |

**Figure 5.**
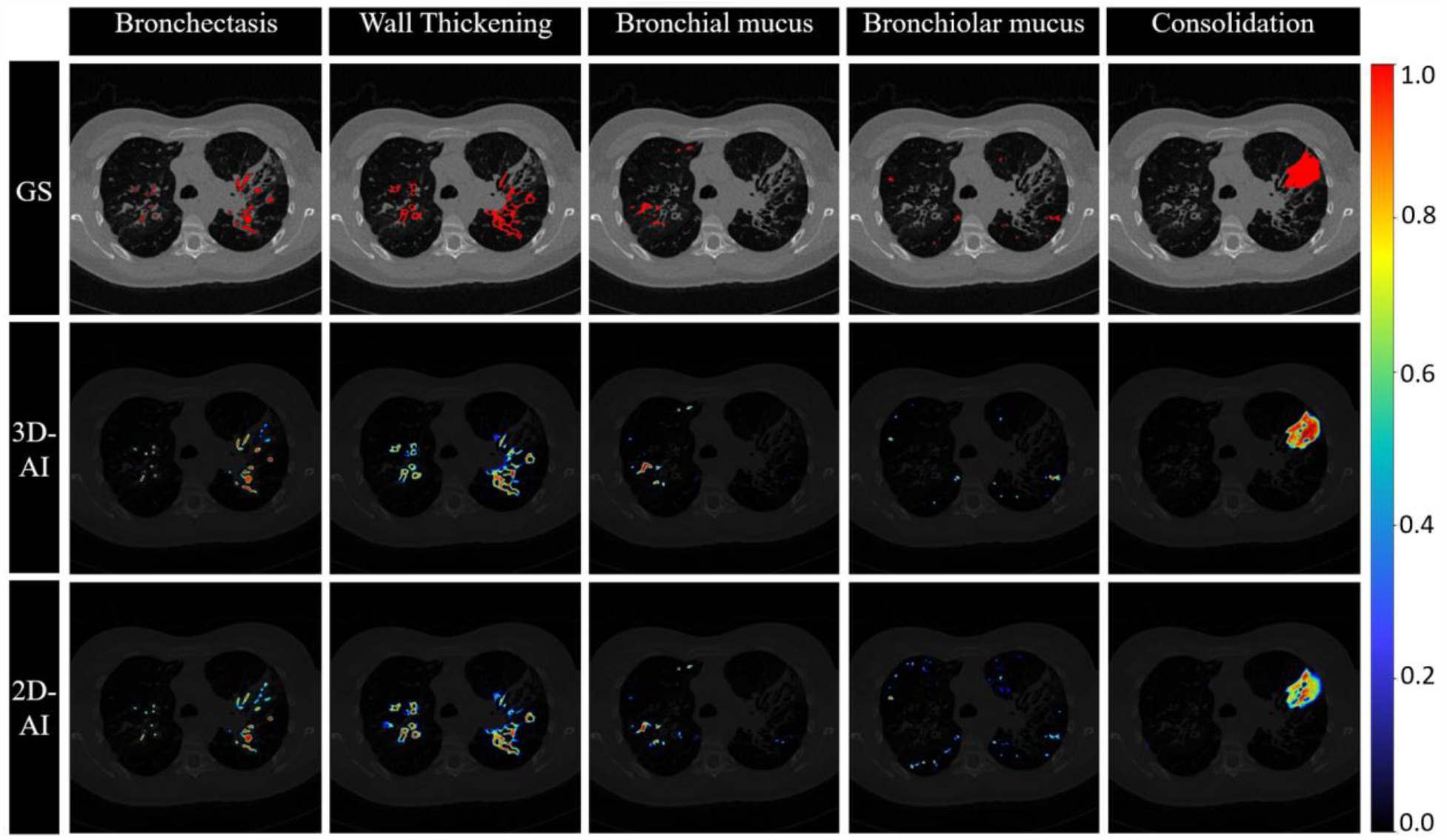
Comparison of CF lesion (Bronchiectasis, Thickening, Bronchial mucus, Bronchiolar mucus, Consolidation) features map. The first row displays the segmentations from the Gold Standard (GS), the second row presents the features map obtained with gradCAM by nnUnet (3D-AI), and the third row depicts the presents the features map obtained produced by nnUnet (2D-AI) gradCAM. The more intense the feature map, the closer the probabilities of the predictions approach 1.

### 2D versus 3D correlations to lung function

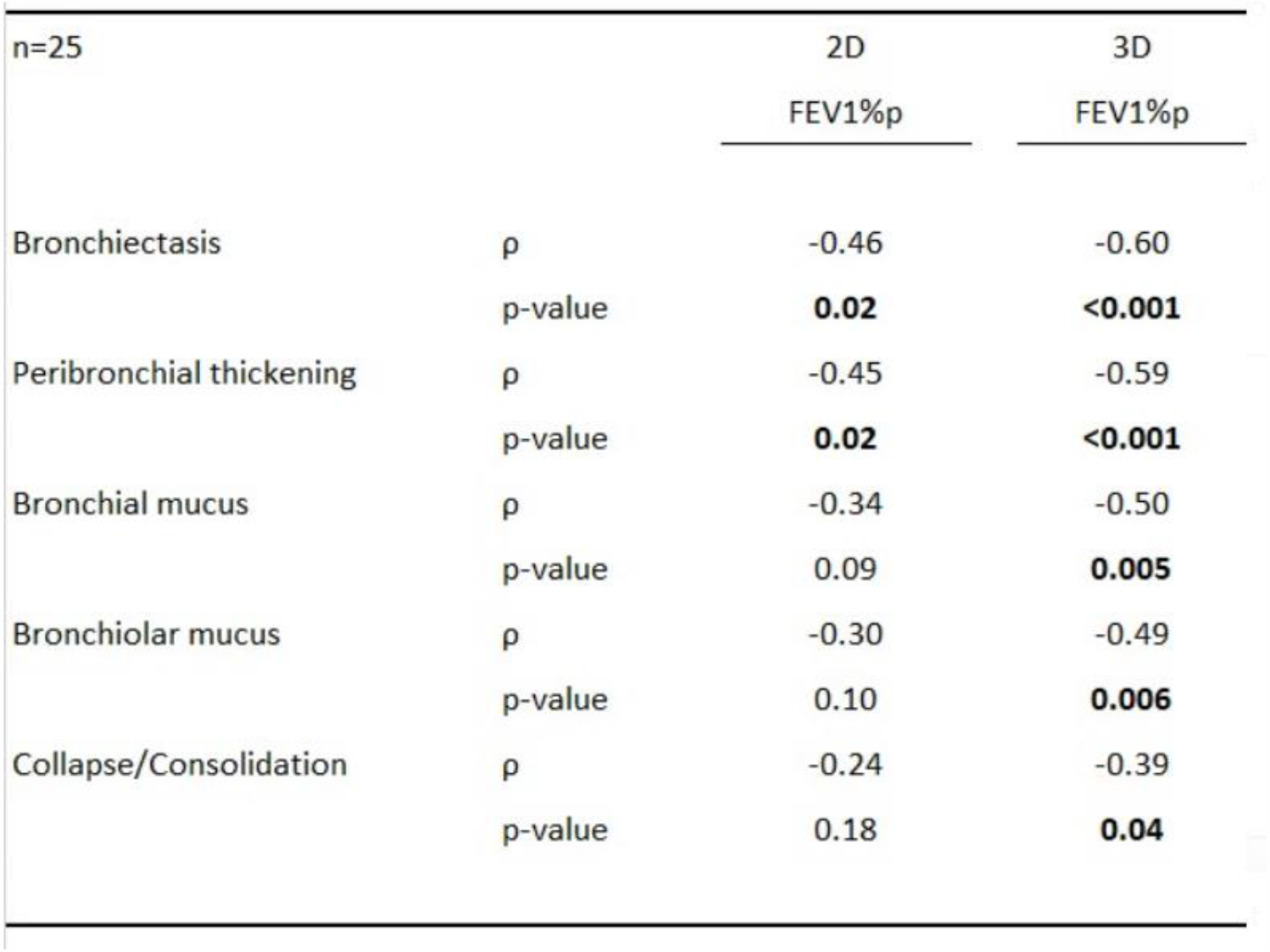

In a test group of 25 patients with CF, there were significant correlations with the forced expiratory volume in 1 second percentage predicted (FEV1%p) in both 2D and 3D regarding Bronchiectasis and Peribronchial thickening volumes. However, the volume segmentations significantly correlated to FEV1% only in 3D regarding bronchial mucus, bronchiolar mucus and collapse/Consolidation.

## DISCUSSION

The 2D and 3D network are posited to have similar aptitude in discerning structures with a significant number of adjacent pixels like bronchectasis, wall thickining and consolidation. However, for more refined structures like bronchial and bronchiolitis mucus, the interlayer spatial information of the 3D training becomes crucial for their detection.

The overlap in identifying different mucus types is understandable due to their semantic closeness. Enhancing model results might have been possible by merging these labels. One potential improvement could have involved employing both parenchymal and mediastinal windows [15] to enrich the network’s knowledge and enhance structure detection.

To avoid perceiving the network as a black box, and in line with recent medical community recommendations [16], model validation via GradCAM was deemed enriching, revealing that the characteristic maps leading to predictions primarily focus on regions of interest and that predictions correlate appropriately with the image structures. Furthermore, the extremely low uncertainty of both models vouched for the consistency and reliability of their predictions.

To date, this research remains the sole endeavor addressing 3D segmentations across five different CF lesions. Existing work [18] focused on assessing dimensions of all visible bronchus-artery (BA) pairs on chest CT and then computing bronchus-vessel ratios to estimate CF progression, a distinct approach from what is proposed here. With the 3D nnUnet, there is potential for monitoring CF progression by comparing the volumes derived from 3D detections of the five CF lesions, offering promising avenues for the management of CF patients.

## CONCLUSION

3D segmentation exhibits superior performance compared to 2D segmentation, particularly in detecting disparate and fine structures. Correlations have been observed between the volumes quantified from 3D segmentation and PFT measurements, underlining the clinical relevance of this method. The implications of these conclusions suggest that there’s room for further exploration in the realm of medical imaging analysis for CF patients.

## Data Availability

All data produced in the present study are available upon reasonable request to the authors

