## Supplemental Method for "2D versus 3D artificial intelligence-driven segmentations of airway alterations in cystic fibrosis: which one is better?"

### Abstract

#### Purpose or Learning Objective

Artificial intelligence (AI) with convolutional neural network allows fully automated detection and segmentation of bronchial changes on CT-scans of cystic fibrosis (CF). However, the superiority of two-dimensional (2D) versus three-dimensional (3D) architectures remains to be explored.

#### Method or Background

CT-scans from fifty CF patients were retrospectively included at two CF reference centers. The nnUnet model was implemented in both 2D and 3D, and trained to segment five structural alterations: bronchiectasis, wall thickening, mucus plugs, bronchiolar impactions and consolidations. A semantic validation was done by using fifty CTs with a five-fold cross validation strategy, by comparing normalized Dice-Sorensen coefficient (DSC) between 2D and 3D architectures, with manual segmentations as Gold Standard.

#### Results or Findings

The 3D nnUnet was found able to segment the five CF main hallmarks such as bronchiectasis, wall thickening, mucus plugs, bronchiolar impactions and consolidations. Metrics obtained with the 3D architecture were superior for mucus plugs, bronchiolar impactions and consolidations ($p<0.001$) but not significantly different for bronchiectasis and wall thickening ($p>0.05$).

**Conclusion**

AI with the 3D-nnUnet model can perform fully automated segmentation of CF-related structural hallmarks on CT scans, and overcome 2D implementation. Non-invasive, holistic 3D quantifications are allowed for promising next clinical applications.

### Supplemental Material

#### 1) Software

The 2D and 3D nnUnet were trained on a system operating on Ubuntu 18.04. The environment was set up with Python 3.9, using~~tilizing~~ PyTorch 2.0 and CUDA 11. The system's hardware was anchored by an Intel Xeon Gold 5217 CPU, featuring 2 physical processors and a total of 32 threads, 30 of which were allocated exclusively to nnU-Net during training. The system was bolstered by 200GB of RAM and employed a Quadro RTX 8000 GPU with 48GB of VRAM.

### 2)

#### 3) Model Architecture:

The nnU-Net [1] is a U-Net based architecture for segmentation, where the encoder extracts essential image features, and the decoder translates these features into a detailed segmentation map.

- 2D Architecture: In the following sections, we explain the components comprising the encoder and decoder of nnU-Net 2D.

1. **Encoder**: The encoder in a nnU-Net 2D leads to extract the features. The features or context extraction block contains 4 layers ordered as follows:
   - i. **2D Convolution**: The 2D convolution operation for a specific output pixel $(x, y)$ is calculated as the sum of element-wise products between the input $Input$ and the convolution kernel $kernel$.

$$(Input * kernel)(x, y) = \sum_i \sum_j Input(x + i, y + j).kernel(i, j)$$

     The 2D convolution operation utilized a kernel size of (3, 3) to process the input data. Additionally, a padding of (1, 1) was applied to the input image. This padding ensured that the convolution operation maintained the spatial dimensions of the input, resulting in an output of the same size.

   - ii. **DropOut**: is a regularization technique used to mitigate overfitting. It involves randomly deactivating a percentage of neurons during training, forcing the network to learn in a more robust and generalizable manner. Regarding our architecture, a dropout rate of 0.3 is employed.

   - iii. **Instance Normalization**: it normalizes activations within each input channel individually, promoting stability and model generalization. Instance normalization is applied to each channel $c$, separately within the feature map. The formula for normalizing an individual pixel $(i, j)$ in channel $c$, is as follows:

$$NormalizedInput_{cij} = \frac{Input_{cij} - \mu_c}{\sqrt{\sigma_c^2 + \varepsilon}}$$

Where $\mu_c$ is the mean of channel $c$, $\sigma_c$ is the variance of channel $c$ .To prevent division by zero when the variance is very small, a small constant called epsilon (ε=$10^{-5}$) is added to the denominator. In this architecture, the momentum is set to $10^{-1}$, and it serves to control the influence of the input $\mu$ and $\sigma$ by weighting them to obtain the current statistics.

iv. **Leaky ReLU (Rectified Linear Unit)**: Leaky ReLU is an activation function that introduces a slight positive slope for negative input values. In the nnU-Net configuration the negative slope $\alpha$ is set to $10^{-2}$. So, for an input $x$ its activation computation is :

$$f(x) = \begin{cases} \alpha x \,, x < 0 \\ x \,, x \geq 0 \end{cases}$$

2. **Decoder**: This part focuses on creating high-resolution feature maps that reflect the spatial details and precise localization of objects or regions of interest in the input. It's responsible for upsampling lower-resolution feature maps generated by the encoder to match the original input resolution.

   This process involves skip connections, which create direct connections between layers at different depths within a network. Those residual connections are inputs to localization block that aimed at identifiying spatial information. The localization block contains 3 layers (2D convolution, instance normalization and leakyRelu). A third block is used to apply deconvolution to increase the spatial resolution of feature maps. It multiplies a transposed sparse kernel by a convolution 2D output.

$$(sparseKernel^t * ConvOutput\,)(x, y) = \sum_i \sum_j sparseKernel^T(x + i, y + j).ConvOutput(i, j)$$

The (Table 1) below summarizes the details of the encoder and decoder parts of the 2D nnU-Net network by listing their types and the number of blocks they contain. The total number of nnU-Net 2D parameters is **41.270.400.**

*Table 1 Details of the blocks comprising the encoder and decoder of the nnU-Net 2D network.*

| | **Type of Blocks** | **Number of unit blocks** | **Layers Details of unit block** |
|---|---|---|---|
| **Encoder** | **Context extraction** | **8*2** | **2D Convolution** |
| | | | **Dropout** |
| | | | **Instance** |

| | | | |
|---|---|---|---|
| | | | **Normalization** |
| | | | **Leaky Relu** |
| **Decoder** | **Features Localization** | **7*2** | **2D Convolution** |
| | | | **Instance Normalization** |
| | | | **Leaky Relu** |
| | **Deconvolution** | **7** | **2D Transpose Convolution** |

- 3D Architecture: For the 3D nnU-Net network, it adheres to the identical architecture, with the exception that the convolution kernels employed are of dimensions (3,3,3). Furthermore, the number of unit blocks differs, which accounts for its lower neuron count compared to the 2D nnU-Net, totaling **30.789.248** parameters. Thus, all the components forming the nnU-Net 3D are enumerated in the subsequent Table 2.

*Table 2 Details of the blocks comprising the encoder and decoder of the nnU-Net 3D network.*

| | **Type of Blocks** | **Number of unit blocks** | **Layers Details of unit block** |
|---|---|---|---|
| **Encoder** | **Context extraction** | **6 * 2** | **3D Convolution** |
| | | | **Dropout** |
| | | | **Instance Normalization** |
| | | | **Leaky Relu** |
| **Decoder** | | **5 * 2** | **3D Convolution** |
| | | | **Instance Normalization** |
| | | | **Leaky Relu** |
| | **Deconvolution** | **5** | **3D Transpose Convolution** |

#### 4) Pre-processing

In order to focus on lesions within the pulmonary parenchyma, the segmentation of the lung envelope was performed using a trained 3D nnU-Net network to recognize it. This segmentation is depicted in Figure 1: on the right, an example of a slice with the envelope is shown (the first input modality of nnU-Net networks), and on the left, the same slice without the lung envelope (the second input modality of nnU-Net networks).

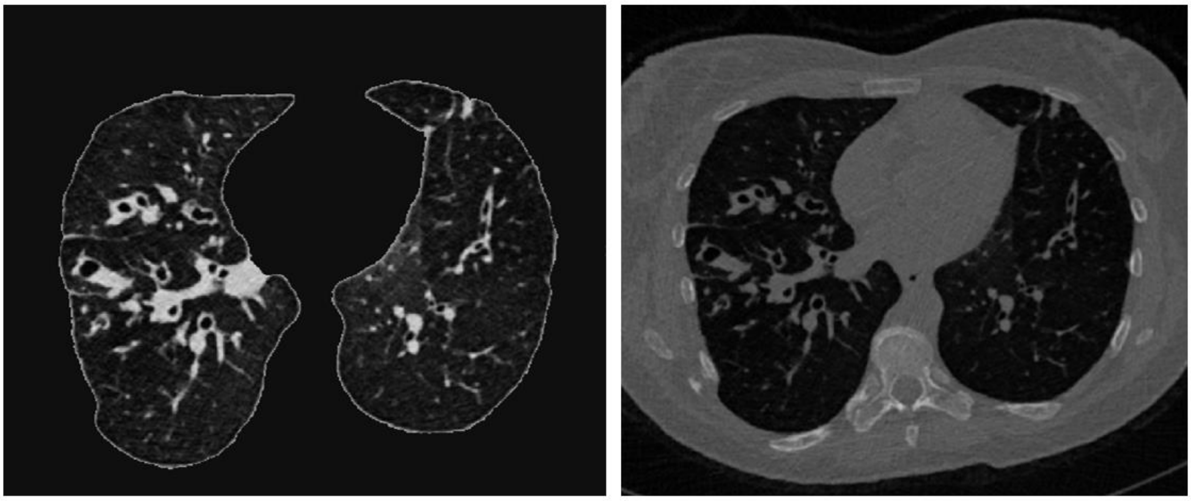

*Figure 1 Two slice inputs of nnU-Net networks, with lung envelope (on the right), without lung envelope (on the left).*

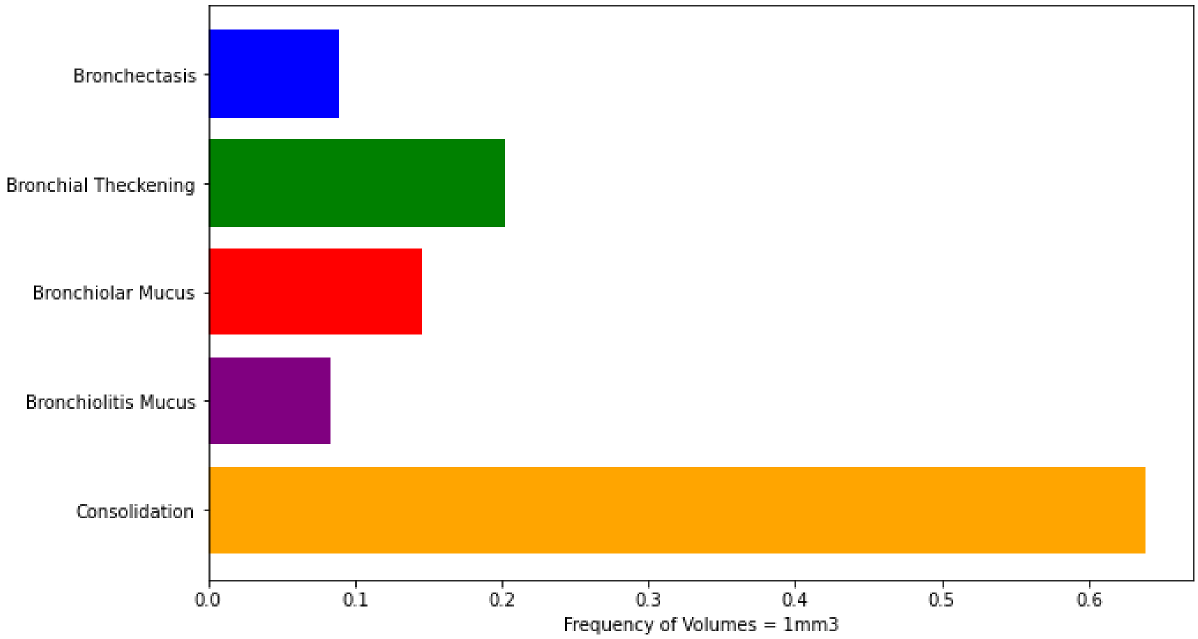


*Figure 2 Frequency of volumes for the 5 pulmonary lesions segmented by nnU-Net*

### 5) Training

The training of both versions of nnU-Net 2D and 3D is carried out in a similar manner: the dataset is divided into 5 folds, with each of the 5 folds serving as a validation set while the remaining 4 folds are used for training the model (i.e., cross-validation). We utilize 50 volumes, so each fold contains 10 volumes with lung envelope and 10 volumes without lung envelope.

In terms of hyperparameters, we mention the following:

- **Loss Function**: it measures the dissimilarity between the model's predictions and the ground truth. In our case, it is a combination of cross-entropy and dice coefficients.
- **Learning Rate (LR):** it determines the step size during parameter optimization. In our case, it starts at $10^{-2}$, and LR is updated every 30 epochs with a minimum threshold of $10^{-6}$.
- **Optimizer**: it is responsible for adjusting the model's parameters to minimize the loss function. nnU-Net uses Stochastic Gradient Descent (SGD) with a Nesterov momentum of 0.99 . SGD iteratively updates the model's parameters to find the minimum of the loss function.
- **Patch**: The patch size is specified for each spatial dimension of the image and varies depending on the nnU-Net version and specific task. In 2D, patches have a size of [512, 512], while in 3D, they have a size of [96, 160, 160].

- **Batch**: It refers to a group of patches that are processed simultaneously by the model during training. In 2D, the batch size is set to 12, while in 3D, it is 2.
- **Epochs**: represent the number of times the entire dataset is processed during training. 1000 epochs are conducted with nnU-Net, and for each epoch, it processes 250 batches. Additionally, the best model is saved every 50 epochs.

The figures below, (Figure 4-8), illustrate the evolution of the Dice score for training (green) and the losses for both training (blue) and validation (red) during the training of nnU-Net 2D. Meanwhile, (Figure 9-13) depict the trend of these measurements (following the same color code) during the training of the 3D version of nnU-Net.

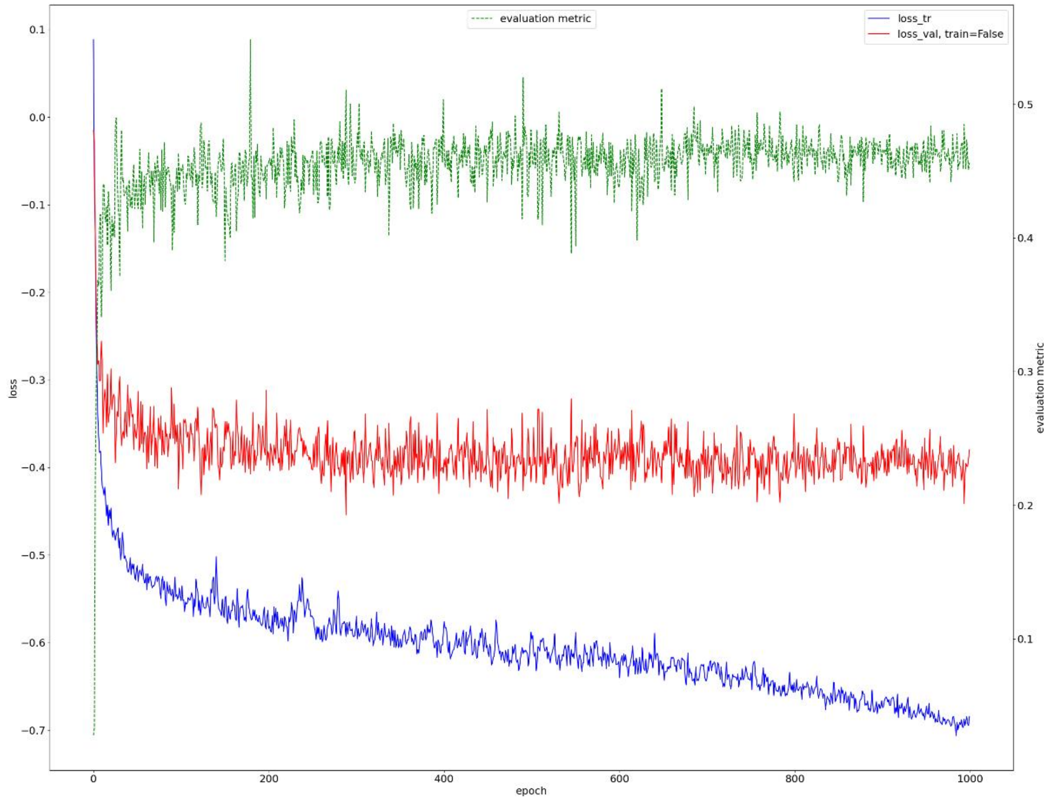


*Figure 4 Measurements of training and validation losses and validation Dice score for Fold 0 during the training of nnU-Net 2D.*

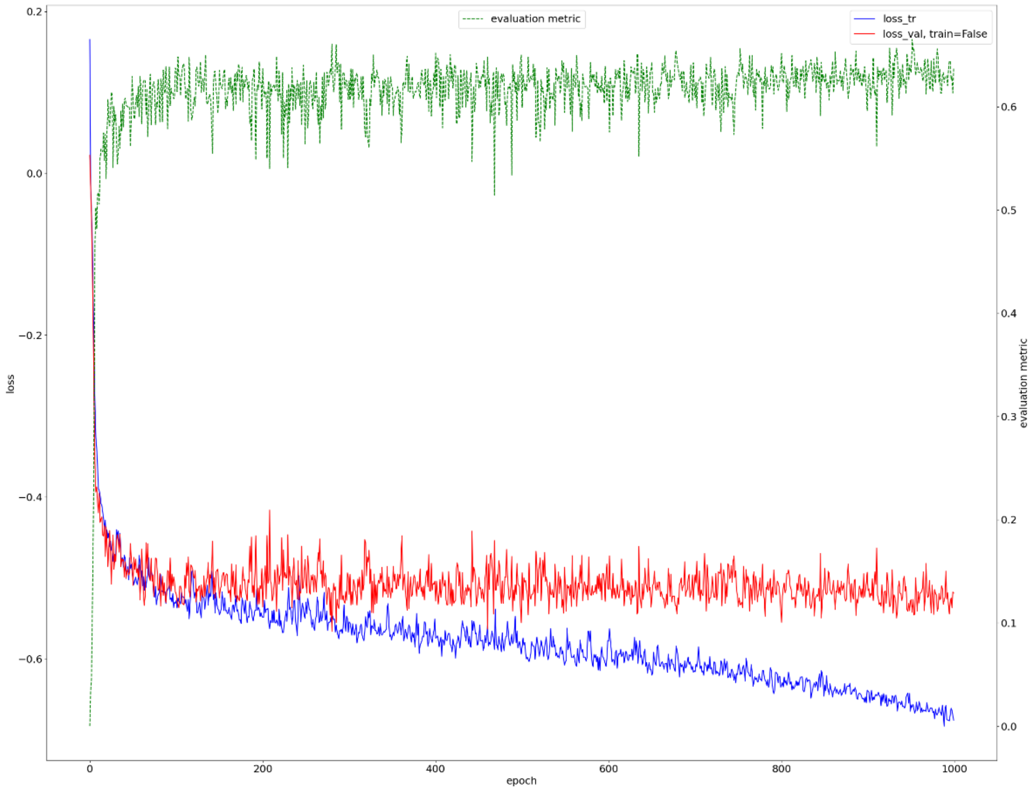


*Figure 5 Measurements of training and validation losses and validation Dice score for Fold 1 during the training of nnU-Net 2D.*

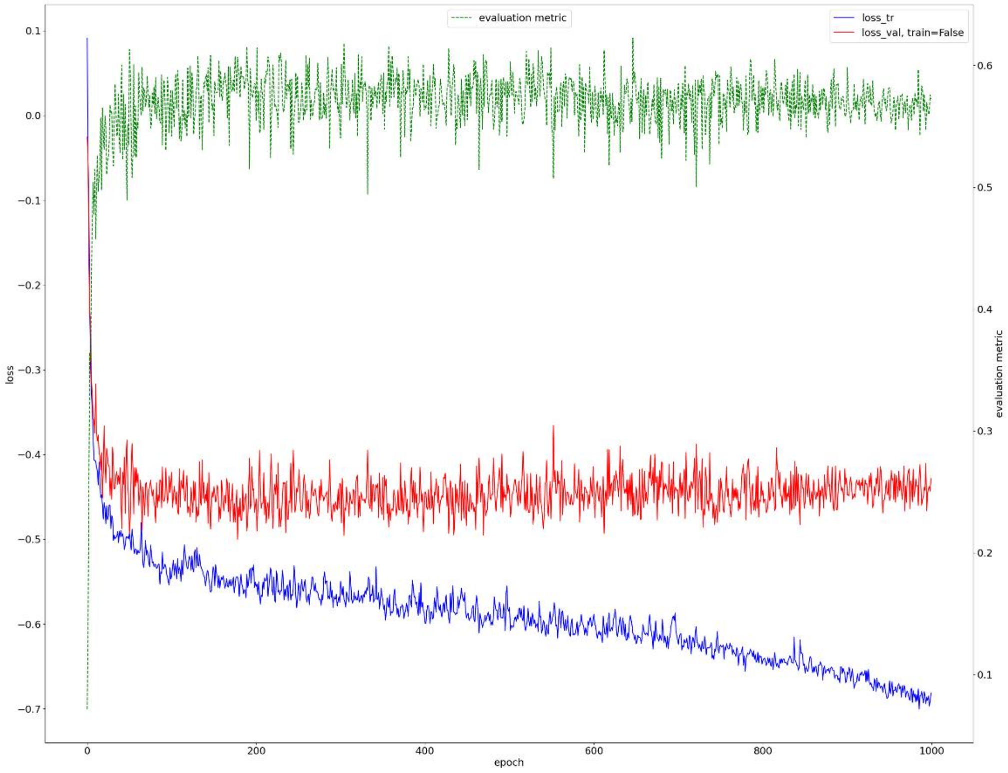


*Figure 6 Measurements of training and validation losses and validation Dice score for Fold 2 during the training of nnU-Net 2D.*

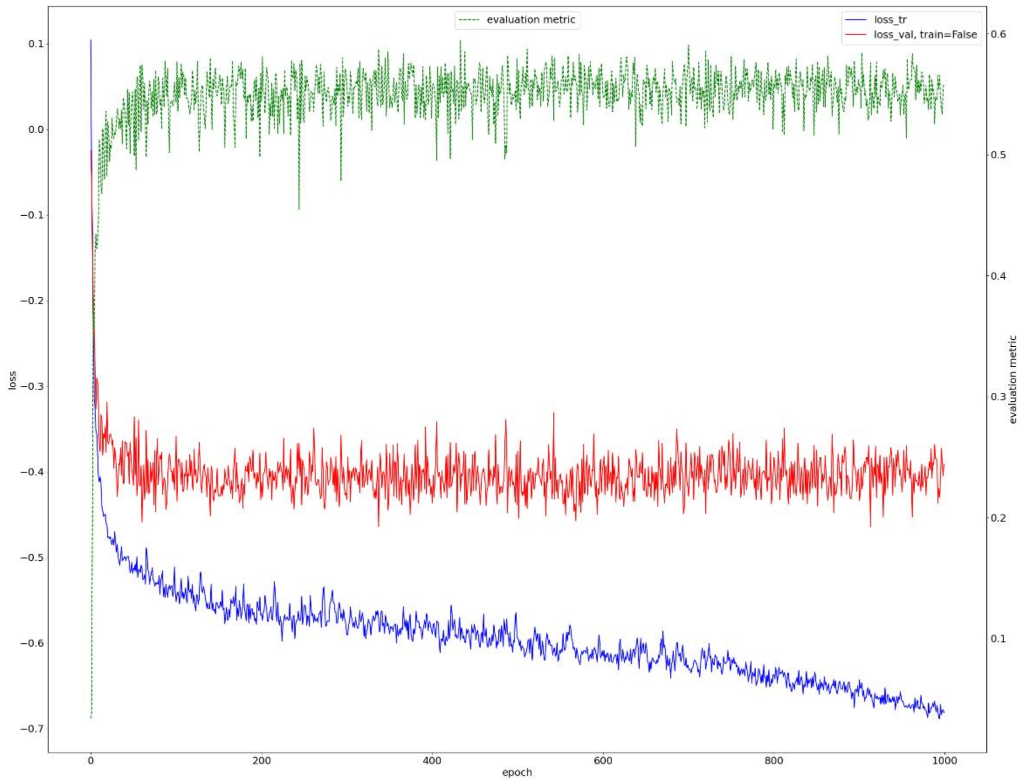


*Figure 7 Measurements of training and validation losses and validation Dice score for Fold 3 during the training of nnU-Net 2D.*

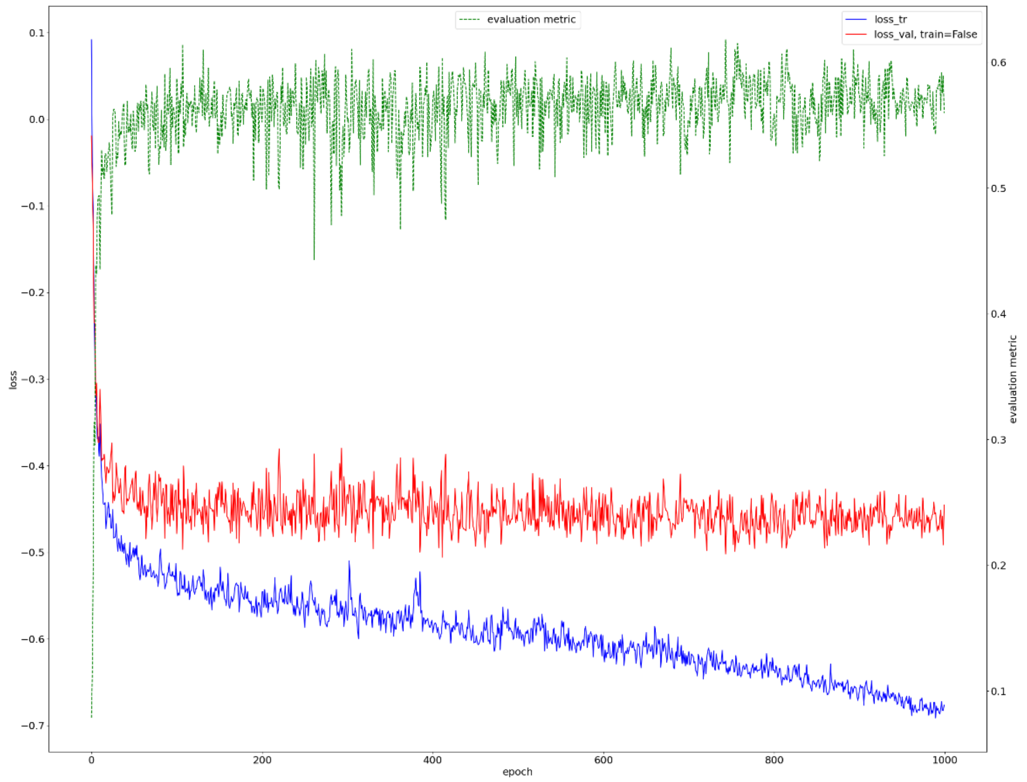


*Figure 8 Measurements of training and validation losses and validation Dice score for Fold 4 during the training of nnU-Net 2D.*

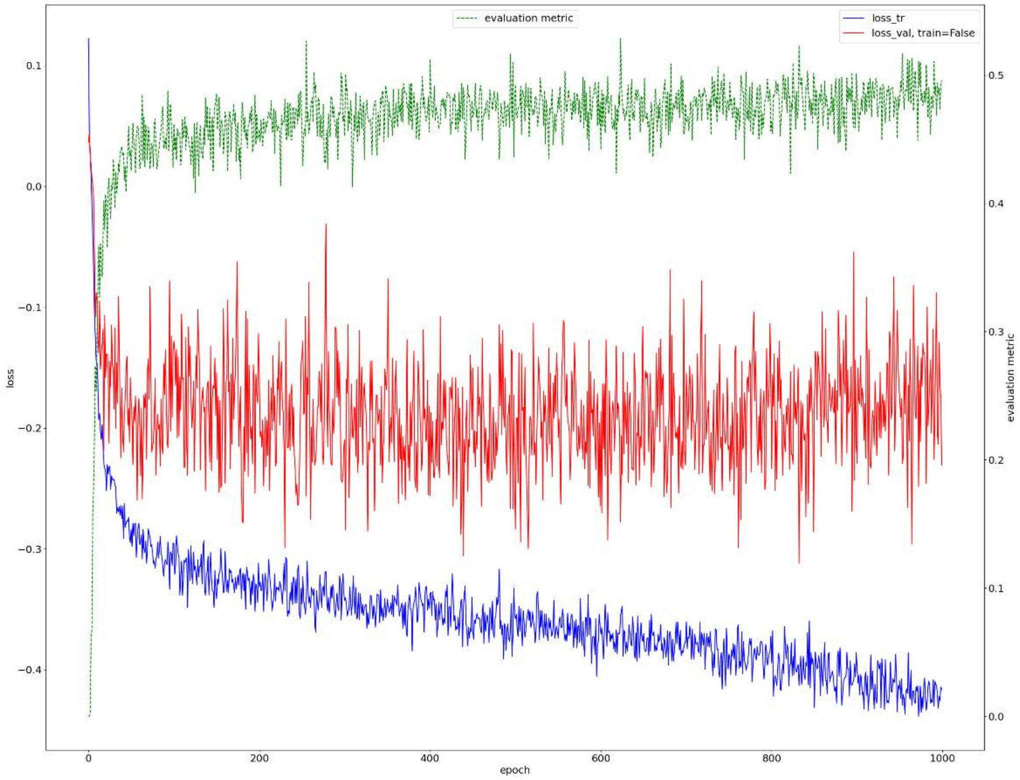


*Figure 9 Measurements of training and validation losses and validation Dice score for Fold 0 during the training of nnU-Net 3D.*

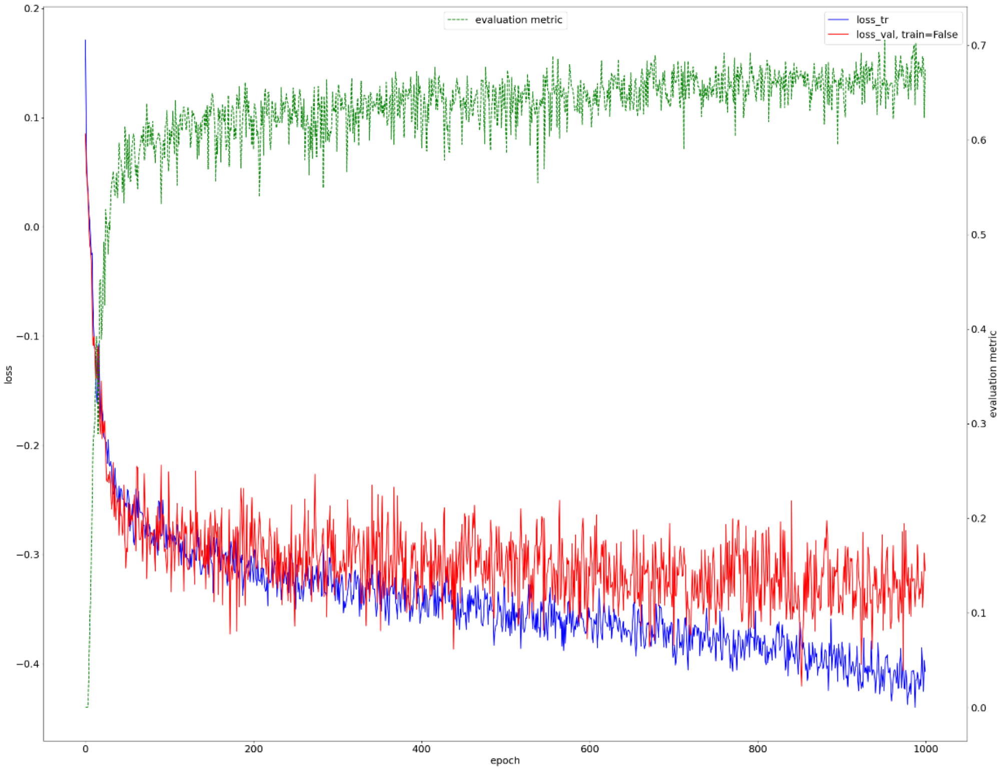


*Figure 10 Measurements of training and validation losses and validation Dice score for Fold 1 during the training of nnU-Net 3D.*

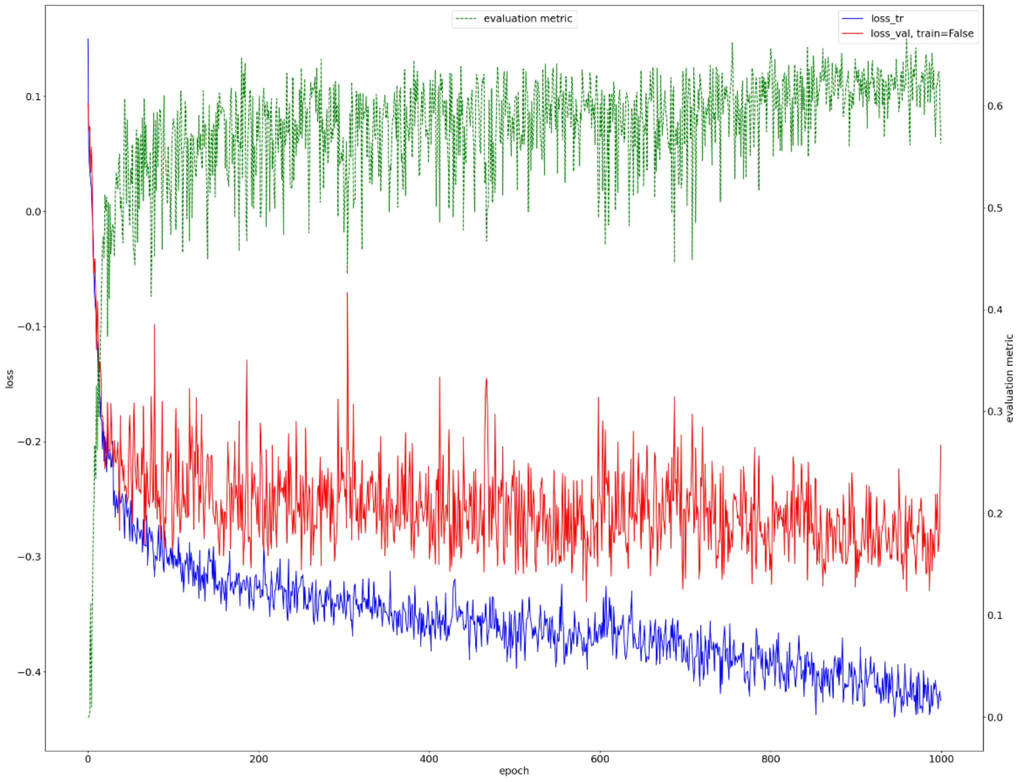


*Figure 11 Measurements of training and validation losses and validation Dice score for Fold 2 during the training of nnU-Net 3D.*

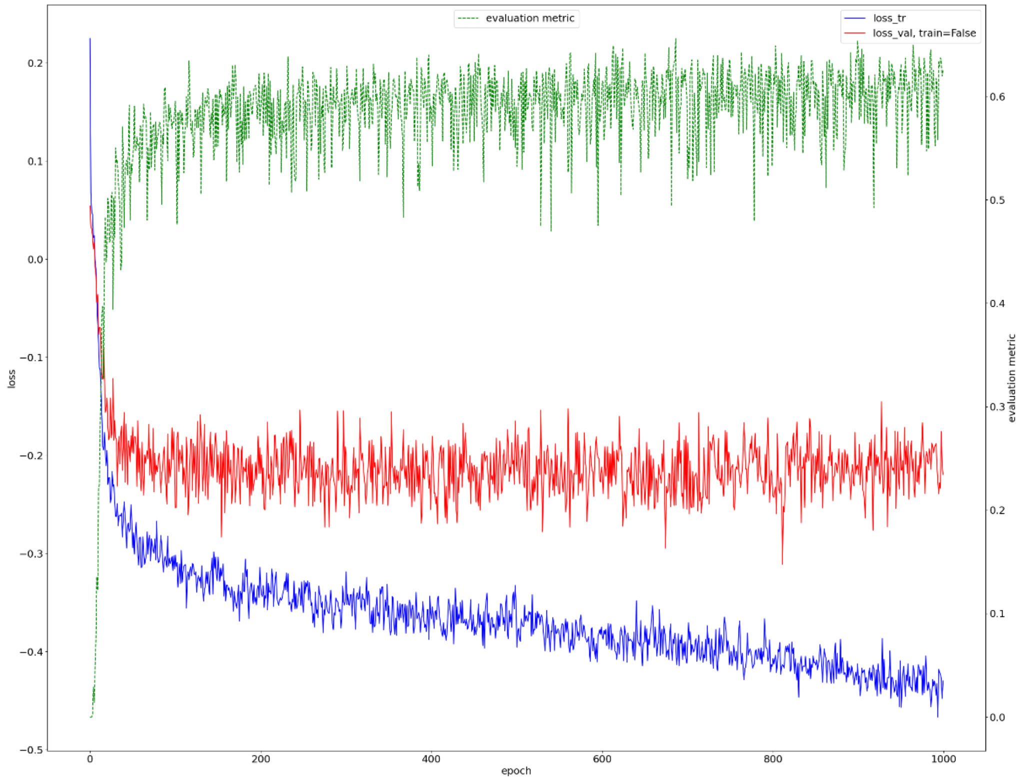


*Figure 12 Measurements of training and validation losses and validation Dice score for Fold 3 during the training of nnU-Net 3D.*

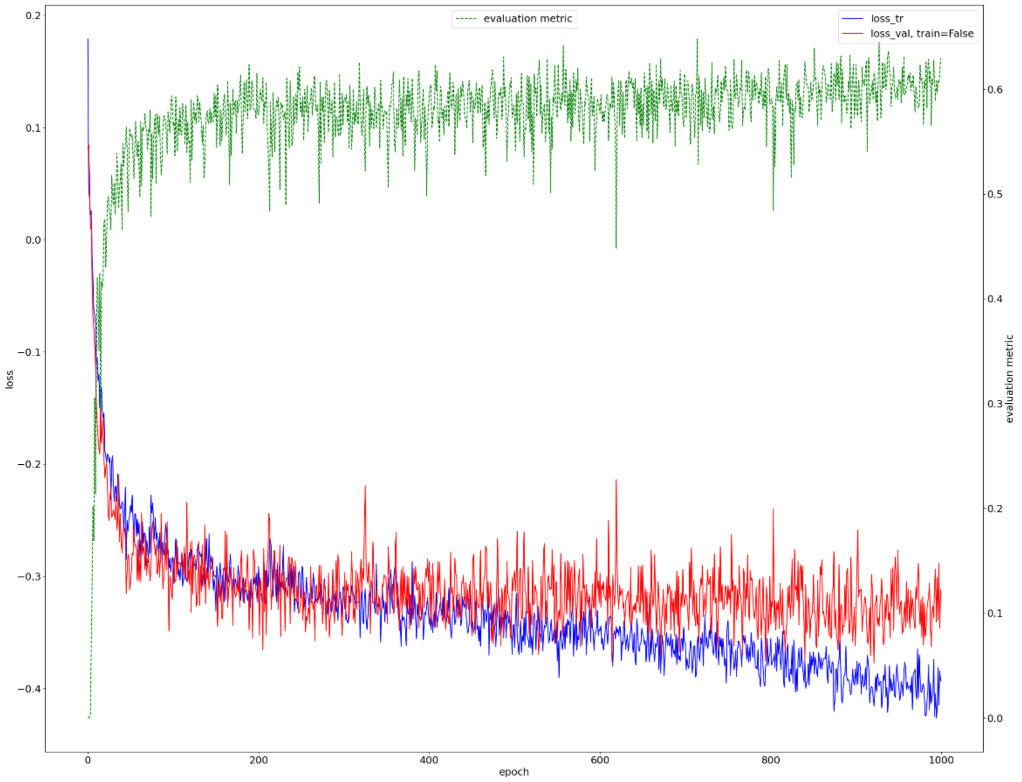


*Figure 13 Measurements of training and validation losses and validation Dice score for Fold 4 during the training of nnU-Net 2D.*

### 6) Evaluation

- **Dice Coefficient:** The Dice coefficient, also known as the F1 score, measures the similarity between two segmentations. It quantifies the overlap between the predicted positive (e.g., segmented region) and true positive (e.g., ground truth) sets.

$$DICE = 2 * (|Predict \cap GroundTruth|) / (|Predict| + |GroundTruth|)$$

With $|Predict \cap GroundTruth|$ the number of pixels present in both the predicted mask and the ground truth mask, $|Predict|$ the total number of pixels in the predicted mask, and $|GroundTruth|$ The total number of pixels in the ground truth mask.

- **Precision:** Precision measures the proportion of true positive pixels among all positive predictions made by the model. It assesses the model's ability to make correct positive pixel predictions.

$$Precision = |Predict \cap GroundTruth| / |Predict|$$

- **Recall (Sensitivity or True Positive Rate):** Recall measures the ability of a model to correctly identify positive instances out of all actual positive instances. It assesses the model's ability to avoid false negatives.

$$Recall = {|Predict\ \cap GroundTruth|}\Big/{|GroundTruth|}$$

- **True Negative Rate (TNR) or Specificity:** TNR or specificity measures the model's ability to correctly identify negative instances out of all actual negative instances.

$$TNR = {|(1 - Predict)\ \cap (1 - GroundTruth)|}\Big/{|(1 - GroundTruth)|}$$

With $|(1 - Predict)\ \cap (1 - GroundTruth)|$ : The number of true negatives (pixels correctly predicted as negatives), and $|(1 - GroundTruth)|$: The total number of negative instances in the ground truth.

- **Area Under the Curve (AUC):** AUC quantifies the model's ability to distinguish between positive and negative labels across different probability thresholds. We focus on one label at a time, setting all other labels to 0. This transforms the problem into binary classification for each label. We then compute the $Recall$ and $TNR$ by varying the prediction threshold for the selected label. The AUC-ROC is determined by integrating the resulting ROC curve.

- **Normalized Surface Dice (NSD):** NSD is closely related to the Dice coefficient, but it provides a nuanced measure of segmentation quality, considering both the volume agreement and the surface smoothness. It measures the similarity between the predicted segmentation and the ground truth while considering a margin around the object's boundary. This margin (3 pixels in our case) accounts for any small variations or uncertainties in the segmentation's border.

$$NSD = {(|Predict_{DB} \cap GT_B| + |GT_{DB} \cap Predict_B|)}\Big/{(|Predic_B| + |GT_B|)}$$

With $|Predict_{DB} \cap GT_B|$ refers to the pixels in the dilated boundary of the prediction and in the ground truth boundary, $|GT_{DB} \cap Predict_B|$ is the pixels present in the dilated boundary of the ground truth and in the prediction boundary, $|GT_B|$ is the ground truth contour, and $|Predict_B|$ is the prediction contour.

The following tables (3-7) display the means results for each label with detailed information for each fold.

- **Confusion Matrix:** It's a table that compares the predicted pixels with the true ground truth pixels for the 5 labels. Each row in the matrix corresponds to a true label, and each column represents a predicted label.

7) **Model Interpretability:** In the realm of clinical decision-making, ensuring model interpretability is paramount for clinicians [2] . This is why two crucial techniques, Grad-CAM and uncertainty calculation, are employed. Grad-CAM visually highlights the

model's decision-making process, offering transparency by showcasing which parts of an image influence its predictions. Simultaneously, calculating uncertainty provides a confidence measure for every prediction.

- **Grad-Cam** [3] **:** is a technique used for visualizing and understanding the decisions made by nnU-Net. It highlights the regions within an input image that the network considers most relevant for making a particular decision. The Grad-CAM map is computed as follows:

*For each label **L** do :*

1. *Isolate L by setting the pixels of all other labels to 0 and the pixels of L to 1*
2. *Feed the input image (with the applied binary mask) through nnU-Net for forward propagation:* $y_L$
3. *Compute the **gradients** of the L score for each feature map* $FM_k$ *in the final convolutional layer (# These feature maps capture various hierarchical and abstract features at different levels of complexity from the input data.)*

$$Gradient\ FM_k = {}^{\partial y_L}/_{\partial FM_k}$$

4. *Considering the overall **importance** of each feature map* $FM_k$ *in prediction by calculating the average of all pixels (#or activations) within a feature map.*

$$\alpha_{FM_k} = \frac{1}{TotalPixel\ FM_k} * \sum_i \sum_j {}^{\partial y_L}/_{\partial FM_k}$$

5. *Get the final Grad-Cam heat map by saving he features that have a **positive influence** on L prediction. ReLU activation is applied on every **weighted map** to save only the positive scores.*

$$GradCAM_L = ReLU\ ( \sum_k \alpha_{FM_k} * FM_k )$$

The figures (14) below depict examples of Grad-CAM computed for three different patients. In each figure, we consider a slice containing the five labels (bronchiectasis (A), bronchial wall thickening (B), bronchiolar mucus (C), bronchiolitis mucus (D), consolidation (E)). Grad-CAMs are calculated for both the 3D (1) and 2D (2) predictions of nnU-Net.

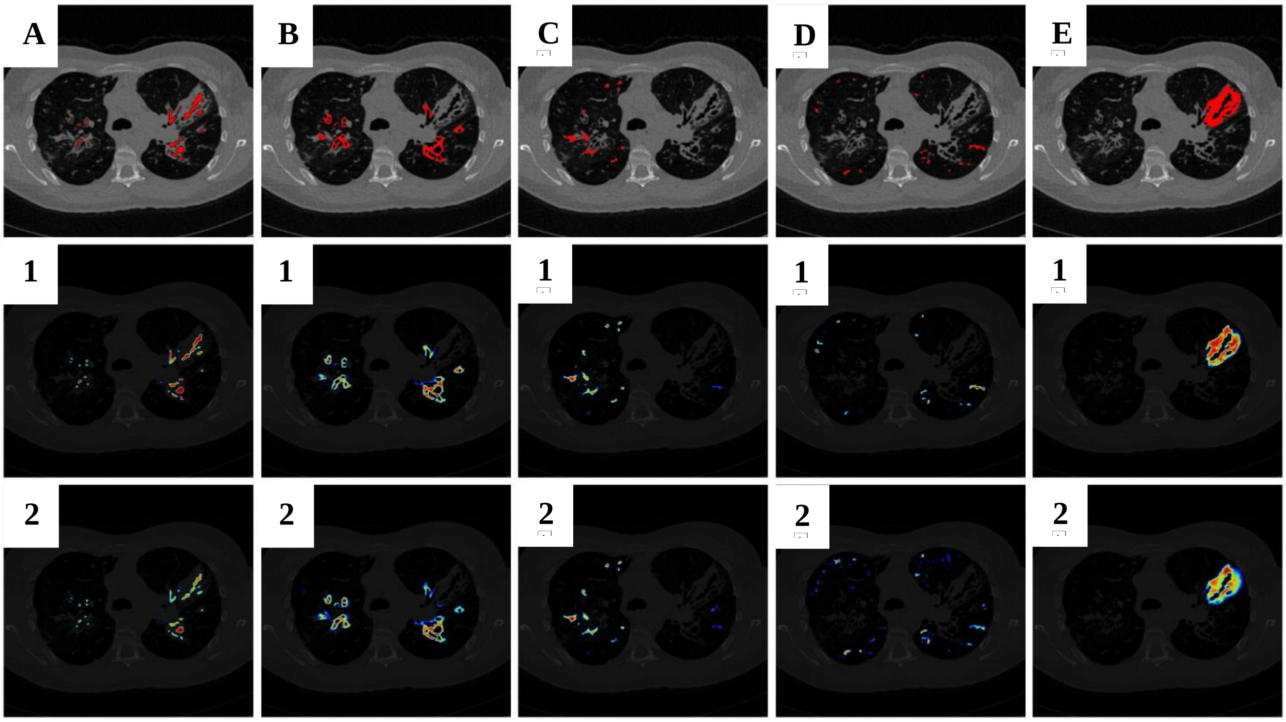
A
B
C
D
E
1
1
1
1
1
2
2
2
2
2

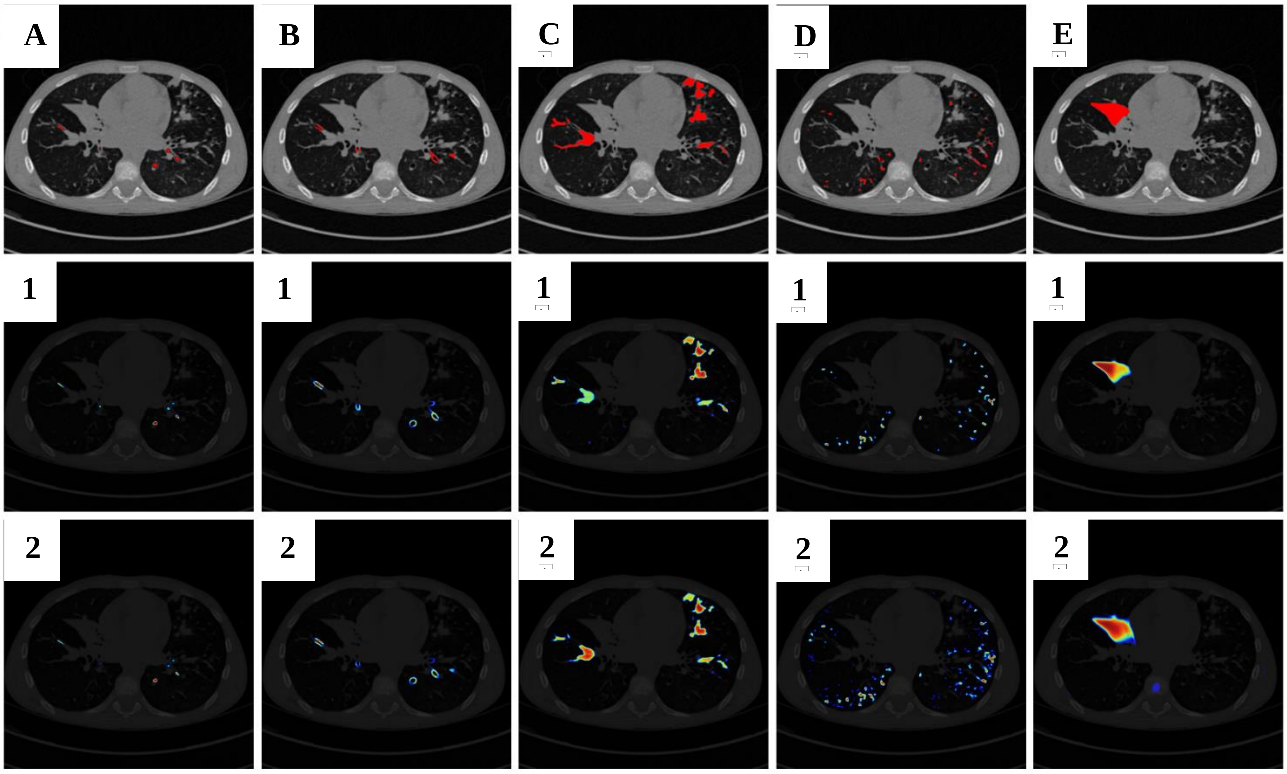
A
B
C
D
E
1
1
1
1
1
2
2
2
2
2

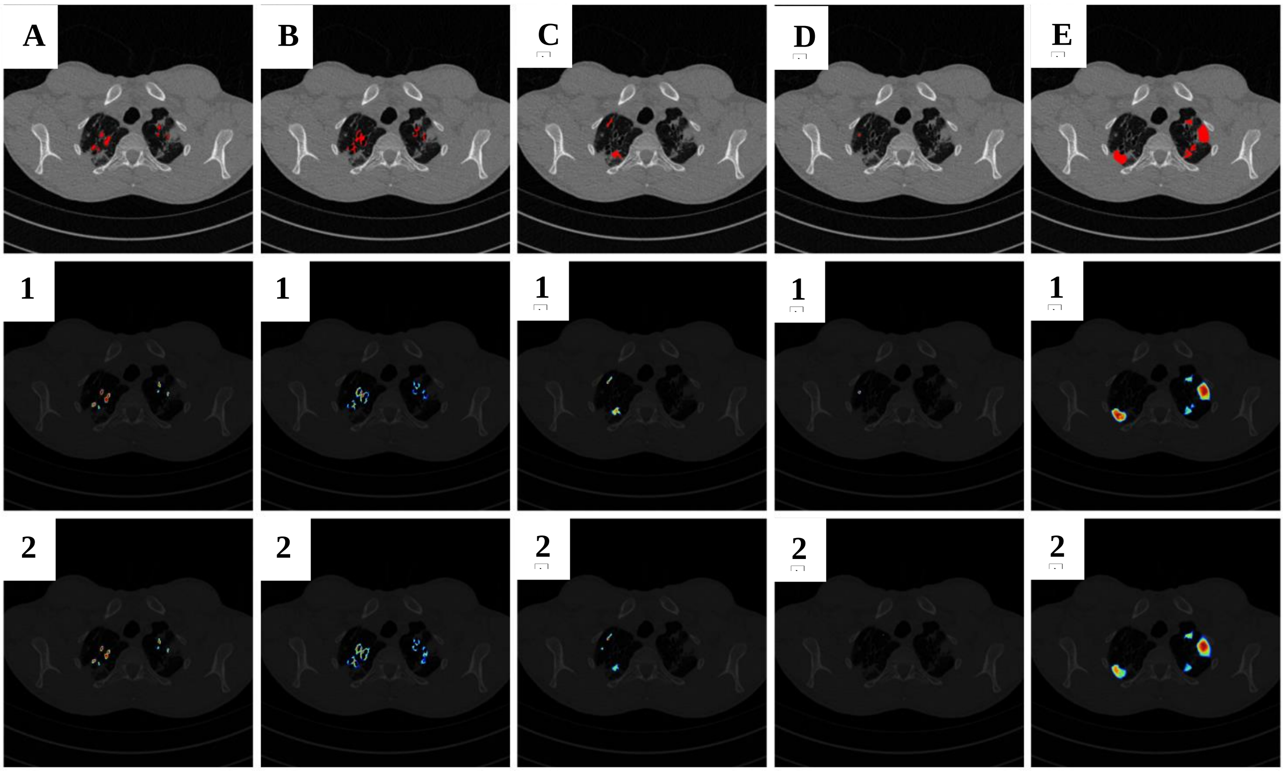


*Figure 14 Example of Grad-CAM calculated for both the 3D and 2D versions of nnU-Net for three different patients.*

- **Uncertainty:** A data-agnostic approach is employed by transforming the network into a Bayesian framework [4], where we treat the weights as distributions rather than fixed values. The uncertainty quantifies the variation in predictions across these model versions to assess the model confidence level. The estimation of the Uncertainty follows these steps:
    1. *Introduce a distribution of weights rather than discrete values: employ 5 times the Monte Carlo Dropout (MC Dropout) by setting the dropout rate to 0.3 during inference. The result is the generation of 5 distinct versions of the network.*
    2. *For each fold $F_k$ (#cross-validation):*
        a. *For each of these network versions $i$ : compute predictions $y_i$ for the input data $x$ from the validation sets.*

$$y_i = Prediction_{nnUnet_i}(x)$$

        b. *Subsequently, Calculate the variance among these five predictions.*

$$VAR\ (F_k) = {\sum(y_i - \overline{y})^2}/{(5-1)}$$

    3. *The model's overall uncertainty is the average of the uncertainties across the 5 folds of the cross-validation.*

$$Uncertainty = \overline{VAR(F_k)}$$

The uncertainty results per fold are listed in the following table 8.

*Table 8 Evaluation results of the uncertainty for nnU-Net 2D and 3D detailed by fold.*

| Label | Modality | Fold 0 | Fold 1 | Fold 2 | Fold 3 | Fold 4 |
|---|---|---|---|---|---|---|
| Bonchectasis | 2D | 0.00043747 | 0.00102918 | 0.000194365 | 0.000210291 | 0.00053768 |
| | 3D | 0.00044165 | 0.00115071 | 0.000208272 | 0.000199593 | 0.00054193 |
| Thickening | 2D | 0.00092219 | 0.00187302 | 0.00053294 | 0.00033216 | 0.00123681 |
| | 3D | 0.00101891 | 0.00168306 | 0.00053256 | 0.00032461 | 0.00123125 |
| Bronchiolar Mucus | 2D | 0.0003214 | 0.00060294 | 0.0004736 | 6.04E-05 | 0.00055551 |
| | 3D | 0.00045454 | 0.00058461 | 0.00058713 | 0.00010545 | 0.00058301 |
| Bronchiolitis Mucus | 2D | 0.00019168 | 0.00026513 | 0.00018466 | 0.00019099 | 0.00039697 |
| | 3D | 0.00041107 | 0.00027858 | 0.00037051 | 0.00022643 | 0.00045317 |
| Consolidation | 2D | 0.00472148 | 0.00034494 | 0.00074111 | 0.0007487 | 0.00012657 |
| | 3D | 0.01113463 | 0.0003614 | 0.00070708 | 0.00059908 | 7.8685E-05 |
